# SARS-CoV-2 Omicron Variant Infection of Individuals with High Titer Neutralizing Antibodies Post-3^rd^ mRNA Vaccine Dose

**DOI:** 10.1101/2022.03.03.22270812

**Authors:** Alexa J. Roeder, Megan A. Koehler, Sergei Svarovsky, Paniz Jasbi, Alim Seit-Nebi, Maria J. Gonzalez-Moa, John Vanderhoof, Davis McKechnie, Baylee A. Edwards, Douglas F. Lake

## Abstract

**Background:** Vaccination with COVID-19 mRNA vaccines prevent hospitalization and severe disease caused by wildtype SARS-CoV-2 and several variants, and likely prevented infection when serum neutralizing antibody (NAb) titers were >1:160. Preventing infection limits viral replication resulting in mutation, which can lead to the emergence of additional variants.

**Methods:** During a longitudinal study to evaluate durability of a three-dose mRNA vaccine regimen (2 primary doses and a booster) using a rapid test that semi-quantitatively measures NAbs, the Omicron variant emerged and quickly spread globally. We evaluated NAb levels measured prior to symptomatic breakthrough infection, in groups infected prior to and after the emergence of Omicron.

**Results:** During the SARS-CoV-2 Delta variant wave, 93% of breakthrough infections in our study occurred when serum NAb titers were <1:80. In contrast, after the emergence of Omicron, study participants with high NAb titers that had received booster vaccine doses became symptomatically infected. NAb titers prior to infection were ≥1:640 in 64% of the Omicron-infected population, ≥1:320 (14%), and ≥1:160 (21%).

**Discussion:** These results indicate that high titers of NAbs elicited by currently available mRNA vaccines do not protect against infection with the Omicron variant, and that mild to moderate symptomatic infections did occur in a vaccinated and boosted population, although did not require hospitalization.

---

SARS-CoV-2 (B.1.1.529) Omicron variant is highly mutated in the receptor-binding domain (RBD) of spike protein and consequently, prone to immune evasion.^1^ Previous reports indicate evasion of vaccine-induced immunity is due in part to a higher degree of neutralizing antibody (NAb) escape compared to NAb escape of previous variants of concern.^2^ While a NAb threshold sufficient to prevent infection and disease remains undefined, particularly regarding the most recent surge in Omicron cases, our findings demonstrate that NAb titers ≥1:640, observed in a majority of our Omicron-infected population, were inadequate for protection from infection and mild to moderate symptomatic disease. Here we report SARS-CoV-2 Omicron variant caused symptomatic infection of 14 individuals who had high titers of neutralizing antibodies after receiving 3^rd^ doses of BNT162b2 or mRNA-1273 vaccines. All individuals were tested on a semi-quantitative rapid test that measures neutralizing antibodies.^3^ Results in **Figure 1** are shown as percent neutralization prior to infection, comparing pre- and post-December 2021 breakthrough infections.

**Figure 1.**
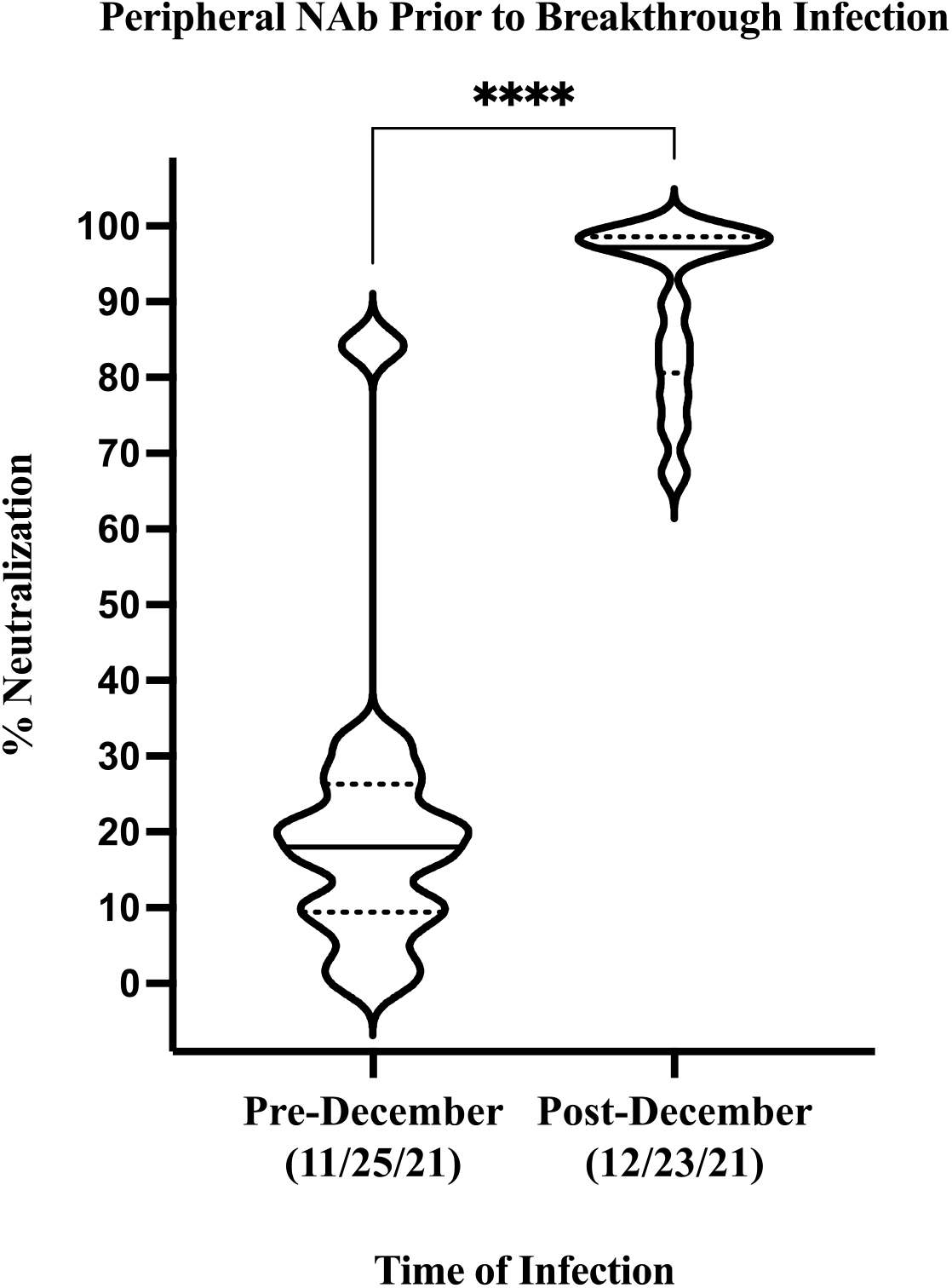
Peripheral NAb prior to breakthrough infection pre- and post-December 2021. NAb data prior to infection are shown as percent neutralization violin plots where solid black lines represent median, and dotted black lines indicate upper and lower quartiles. Significance was assessed using a non-parametric Mann-Whitney test to evaluate mean rank between groups with a two-tailed *p*<0.0001 and 95% confidence interval. A single outlier in the pre-December 2021 group had 84% neutralization prior to infection, however, was on the 2^nd^ week of a 40 mg/day prednisone taper at the time of infection. Demographic information pertaining to individuals in both groups are detailed in **Supplementary Table S1**.

Prior to December of 2021 and the Omicron surge in the USA, we observed only 14 PCR-confirmed breakthrough infections in our study population of 269 mRNA vaccine recipients. Thirteen of those breakthrough infections occurred when NAb levels had declined to titers <1:80 with an average of 16% neutralization (range 0-84%, median 17%). Only one individual in the pre-December 2021 breakthrough population had a NAb titer >1:80 (**Supplementary Figure S1**). Conversely, 14 individuals that had high NAb titers (≥1:640 [n=9], ≥1:320 [n=2] and ≥1:160 [n=3], average 90% neutralization) after receiving a 3^rd^ mRNA vaccine dose became symptomatically infected after Omicron became the dominant variant in circulation,^4^ (range 67-99% neutralization, median 97%) (**Figure 1**). Population demographics, vaccination data, and time between NAb and PCR testing are shown in the Supplementary Appendix (**Table S1**).

Although one limitation of this study is small sample size, our results are the first to show that high neutralizing antibody titers induced by a 3rd dose of currently available mRNA vaccines do not protect against symptomatic breakthrough infections with the SARS-CoV-2 Omicron variant. However, other reports have also observed symptomatic breakthrough infection in individuals with mRNA vaccine booster doses.^5^ It is important to note that all Omicron infections described in this study self-reported symptoms as mild to moderate, such as upper respiratory congestion, sore throat, cough, headache, myalgia, lymphadenopathy, and fever (<39.4°C). Although a higher NAb titer is seemingly less protective against Omicron (B.1.1.529) than with previous variants of concern, our data support other reports that current vaccines remain highly effective at preventing severe disease.^6,7^ Most notably, our data demonstrate that even those at the highest NAb titers detected by our rapid test (≥1:640) remain susceptible to symptomatic Omicron infection and are presumably capable of transmitting virus.

## Data Availability

All data produced in the present work are contained in the manuscript and supplementary materials. Additional data not included are available upon reasonable request to the corresponding author.

