## Supplementary Appendix for "SARS-CoV-2 Omicron Variant Infection of Individuals with High Titer Neutralizing Antibodies Post-3^rd^ mRNA Vaccine Dose"

This appendix has been provided by authors to give readers additional information about the work.

### A) Pre-December 2021

| Age | Sex | 1st/2nd Dose mRNA Vaccine | Date of 2nd Dose | Pre-Infection NAb Test | Pre-Infection % Neut. (NAb Titer) | Date of PCR+ | NAb Test to PCR+ (days) |
|---|---|---|---|---|---|---|---|
| 46-50 | M | BNT162b2 | Mar-2021 | Sep-2021 | 84.2 (>1:320) | Sep-2021 | 8 |
| 31-35 | M | BNT162b2 | Jan-2021 | Apr-2021 | 32.3 (<1:80) | Oct-2021 | 187 |
| 26-30 | F | BNT162b2 | Jan-2021 | Jul-2021 | 28.5 (<1:80) | Aug-2021 | 18 |
| 61-65 | M | BNT162b2 | Jan-2021 | Aug-2021 | 25.6 (<1:80) | Sep-2021 | 20 |
| 76-80 | F | BNT162b2 | Jan-2021 | May-2021 | 21.3 (<1:80) | Jul-2021 | 61 |
| 31-35 | F | mRNA-1273 | Mar-2021 | Sep-2021 | 20.8 (<1:80) | Nov-2021 | 40 |
| 51-55 | M | BNT162b2 | Feb-2021 | Sep-2021 | 19.4 (<1:80) | Sep-2021 | 8 |
| 51-55 | F | mRNA-1273 | Feb-2021 | Aug-2021 | 16.6 (<1:80) | Sep-2021 | 21 |
| 76-80 | M | BNT162b2 | Feb-2021 | May-2021 | 16.3 (<1:80) | Jul-2021 | 53 |
| 51-55 | F | mRNA-1273 | Feb-2021 | Sep-2021 | 10.8 (<1:40) | Nov-2021 | 69 |
| 56-60 | M | BNT162b2 | Feb-2021 | Oct-2021 | 10.1 (<1:40) | Oct-2021 | 3 |
| 81-85 | F | BNT162b2 | Feb-2021 | Sep-2021 | 7.3 (<1:40) | Oct-2021 | 23 |
| 46-50 | F | BNT162b2 | Feb-2021 | Oct-2021 | 2.9 (<1:40) | Nov-2021 | 43 |
| 41-45 | F | BNT162b2 | Mar-2021 | Aug-2021 | 0.0 (<1:40) | Sep-2021 | 17 |

### B) Post-December 2021

| Age | Sex | 1st/2nd Dose mRNA Vaccine | 3rd Dose mRNA Vaccine | Date of 3rd Dose | Pre-Infection NAb Test | Pre- Infection % Neut. (NAb Titer) | Date of PCR+ | NAb Test to PCR+ (days) |
|---|---|---|---|---|---|---|---|---|
| 31-35 | M | mRNA-1273 | mRNA-1273 | Nov-2021 | Dec-2021 | 98.9 (>1:640) | Jan-2022 | 26 |
| 61-65 | F | BNT162b2 | BNT162b2 | Oct-2021 | Dec-2021 | 98.8 (>1:640) | Dec-2021 | 19 |
| 56-60 | F | BNT162b2 | BNT162b2 | Oct-2021 | Jan-2022 | 98.8 (>1:640) | Jan-2022 | 17 |
| 46-50 | F | BNT162b2 | mRNA-1273 | Sep-2021 | Dec-2021 | 98.5 (>1:640) | Jan-2022 | 34 |
| 51-55 | F | BNT162b2 | mRNA-1273 | Oct-2021 | Dec-2021 | 98.5 (>1:640) | Jan-2022 | 27 |
| 51-55 | F | BNT162b2 | BNT162b2 | Oct-2021 | Dec-2021 | 98.4 (>1:640) | Jan-2022 | 20 |
| 36-40 | F | BNT162b2 | mRNA-1273 | Nov-2021 | Nov-2021 | 97.4 (>1:640) | Jan-2022 | 60 |
| 46-50 | M | BNT162b2 | mRNA-1273 | Aug-2021 | Nov-2021 | 96.9 (>1:640) | Jan-2022 | 34 |
| 46-50 | M | BNT162b2 | BNT162b2 | Oct-2021 | Dec-2021 | 89.7 (>1:640) | Jan-2022 | 27 |
| 41-45 | F | BNT162b2 | BNT162b2 | Oct-2021 | Dec-2021 | 85.0 (>1:320) | Jan-2022 | 25 |
| 66-70 | M | mRNA-1273 | mRNA-1273 | Oct-2021 | Dec-2021 | 81.6 (>1:320) | Jan-2022 | 30 |
| 61-65 | M | BNT162b2 | BNT162b2 | Aug-2021 | Dec-2021 | 77.6 (>1:160) | Dec-2021 | 18 |
| 41-45 | F | BNT162b2 | BNT162b2 | Oct-2021 | Dec-2021 | 73.4 (>1:160) | Jan-2022 | 24 |
| 36-40 | F | mRNA-1273 | mRNA-1273 | Sep-2021 | Jan-2022 | 67.4 (>1:160) | Jan-2022 | 6 |

**Supplementary Table S1. (A)** Pre- and **(B)** Post-December 2021 population demographics, vaccination status, and NAb result preceding infection. Age, sex, 1st/2nd vaccine dose and date, 3rd dose vaccine and date (if applicable), date of last NAb test prior to infection, pre-infection NAb result, date of positive PCR for SARS-CoV-2 detection, and days between NAb and PCR dates are detailed in this table. Pre-infection NAb test result is shown as % neutralization, calculated using the limit of detection our rapid NAb lateral flow as described previously.[1] Average age of the pre- and post-December 2021 populations were 54 and 49 (median 55 and 49.5), respectively. Populations in (A) and (B) were 57% and 64% female, respectively.

Supplementary Figure S1 is provided to help orient readers with respect to conversion between units described in the main text, such as NAb LFA test line density, percent neutralization, and NAb titer.

| | | | | | | |
|---|---|---|---|---|---|---|
| **NAb LFA Test Image** | 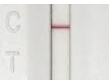 | 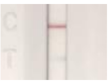 | 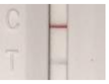 | 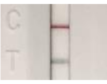 | 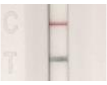 | 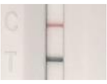 |
| **Test Line Density Units (thousands)** | 10-99 | 100-199 | 200-369 | 370-599 | 600-799 | 800-1000 |
| **Neutralization (%)** | 99-90 | 89-80 | 79-61 | 60-36 | 35-15 | ≤15 |
| **NAb Titer** | <1:1280<br>≥1:640 | <1:640<br>≥1:320 | <1:320<br>≥1:160 | <1:160<br>≥1:80 | <1:80<br>≥1:40 | ≤1:40 |

**Supplementary Figure S1.** Neutralizing antibody later flow assay (NAb LFA) measured in density units corresponds to percent neutralization and NAb titer. Validation of the rapid NAb test using gold standard focus reduction neutralization test ($FRNT_{50}$) with authentic SARS-CoV-2 Wuhan isolate and correlation/regression analyses are previously described.[1,2]
